# An integrated lab-on-a-chip device for RNA extraction, amplification and CRISPR-Cas12a-assisted detection for COVID-19 screening in resource-limited settings

**DOI:** 10.1101/2022.01.06.22268835

**Authors:** Bongkot Ngamsom, Alexander Iles, Moses Kamita, Racheal Kimani, Pablo Rodriguez-Mateos, Mary Mungai, Charlotte E. Dyer, Cheryl Walter, Jesse Gitaka, Nicole Pamme

## Abstract

In response to the ongoing COVID-19 pandemic and disparities of vaccination coverage in low- and middle-income countries, it is vital to adopt a widespread testing and screening programme, combined with contact tracing, to monitor and effectively control the infection dispersion in areas where medical resources are limited. This work presents a lab-on-a-chip platform, namely “IFAST-CRISPR”, as an affordable, rapid and high-precision molecular diagnostic means for SARS-CoV-2 detection. The herein proposed “sample-to-answer” platform integrates RNA extraction, amplification and CRISPR-Cas-based detection with lateral flow readout in one device. The microscale dimensions of the device containing immiscible liquids, coupled with the use of silica paramagnetic beads and GuHCl, streamline sample preparation, including RNA concentration, extraction and purification, in 15 min with minimal hands-on steps. By combining RT-LAMP with CRISPR-Cas12 assays targeting the nucleoprotein (N) gene, visual identification of ≥ 470 copies mL^-1^ genomic SARS-CoV-2 samples was achieved in 45 min, with no cross-reactivity towards HCoV-OC43 nor H1N1. On-chip assays showed the ability to isolate and detect SARS-CoV-2 from 1,000 genome copies mL^-1^ of replication-deficient viral particles in 1 h. This simple, affordable and integrated platform demonstrated a visual, faster, and yet specificity and sensitivity-comparable alternative to the costly gold-standard RT-PCR assay, requiring only a simple heating source. Further investigations on multiplexing and direct interfacing of the accessible Swan-brand cigarette filter for saliva sample collection could provide a complete work flow for COVID-19 diagnostics from saliva samples suitable for low-resource settings.

## 1. Introduction

Since the first case of coronavirus disease 2019 (COVID-19) caused by Severe Acute Respiratory Syndrome Coronavirus 2 (SARS-CoV-2) was reported in Wuhan, China in December 2019, more than 260 million cases and 5.2 million deaths have been reported as of December 7, 2021 (1). Many cases of severe illness and death associated with COVID-19 infections have been prevented in countries where there has been effective implementation of vaccine programmes with ca. 42% of the world population fully vaccinated (data as of December 2021, (2)). However, disparities in vaccination coverage are an ongoing issue, especially in low- and middle-income countries where, on average, only 6.3% of the population have received one dose. In particular, instances of less than 2% coverage have been reported in more than 20 African countries (3). Considering the rush for apportionment of the global supply of vaccines between nations, and the inequitable distribution between high and poor-income countries, and emergence of highly transmissible omicron variant (4), increased testing and screening with contact tracing are paramount for infection control in areas where resources are limited.

The gold standard for COVID-19 diagnostics detects the presence of viral RNA from respiratory specimens using real-time reverse transcription-polymerase chain reaction (RT-qPCR) (5-7). The analytical limits of detection of RT-qPCR are usually ca. 10^3^ viral RNA copies mL^-1^ with 24-48 h turnaround time (6, 8). Despite the high sensitivity and specificity for disease diagnosis, the method relies on a highly skilled operator using expensive instrumentation, reducing its applicability for widespread application (6, 9, 10).

Several point-of-care (POC) RNA detection technologies, requiring no special instruments, employ simultaneous reverse transcription (RT) and isothermal amplification steps such as loop-mediated isothermal amplification (LAMP, (11, 12)) or recombinase polymerase amplification (RPA, (13)). Although displaying high sensitivity, RT-LAMP and RT-RPA can suffer from nonspecific amplification under isothermal conditions, leading to false-positive results (14, 15). Other automated commercially available systems such as Cepheid GeneXpert, Roche Cobas and Abbot ID Now offer the ease-of-use aspect with minimal hands-on steps with ≤ 1 h sample-to-result; however, expensive specialized instrumentation is still required for each system (16).

An alternative to nucleic acid amplification tests, antigen lateral flow assays detect the presence of the virus through specific nucleocapsid proteins of SARS-CoV-2. They require no RNA purification step, and can operate at ambient temperatures with ≤ 30 min turnaround time, making them appealing within POC settings. However, the lack of exponential amplification can hinder the detection limit with a sensitivity of the assays typically being three orders of magnitude lower than the gold standard RT-qPCR (17).

Clustered regularly interspaced short palindromic repeats (CRISPR)-based diagnostic methods utilize the collateral cleavage activity of bystander nucleic acid probes of RNA-guided CRISPR-associated 12/13 (Cas12/13) nucleases (15, 18-21). In combination with RT-LAMP or RT-RPA isothermal amplification methods, CRISPR-Cas-assisted SARS-CoV-2 detection assays are considered as transformative methods for POC COVID-19 diagnostics (15, 21-25). The highly sensitive and specific nature of CRISPR diagnostic methods is attributed to the sequence specificity required for both nucleic acid amplification step and the CRISPR-Cas detection step (15). Readout of Cas-mediated nucleic acid probe cleavage can either be by fluorescence detection or via lateral flow strip, both of which can be applied for POC purposes (21, 26). The original DETECTR-based (21), and SHERLOCK-based (15) detections involved multiple manual steps and operated with already extracted RNA, thereby limiting their applicability for POC use (27). Although the more recent STOPCovid.v2 assay included a benchtop magnetic-based RNA extraction prior to the one-step CRISPR-Cas12b-assisted RT-LAMP (28), the assay still requires multi-step manual operation.

Numerous efforts have been made in developing fully integrated, affordable, sensitive, specific, rapid and robust platforms for POC COVID-19 diagnostics exploiting microfluidics or ‘lab-on-a-chip’ technologies (16, 29). Such platforms permit viral lysis, RNA extraction, nucleic acid amplification and detection on a single device, allowing more applicable diagnostics within resource-limited settings (27, 30). Immiscible filtration assisted by surface tension (IFAST) is a microfluidic technique which exploits the surface tension properties of immiscible liquids and microscale dimensions to facilitate side-by-side compartmentation of immiscible liquids, enabling multiple and sequential steps to be performed in one device (31-33). The use of suitable functionalized magnetic particles allows rapid isolation and purification of a magnetically responsive analyte from complex matrices (32, 34-39).

Here, IFAST and the recent developments in CRISPR-Cas12-based sensing applied to SARS-CoV-2 detection are combined to develop a cost-effective, sensitive, target-specific and fully integrated device for COVID-19 diagnostics, namely ‘IFAST-CRISPR’, Fig. 1A. The device was designed for streamlined sample preparation to provide rapid isolation and concentration of RNA directly from nasopharyngeal swabs or saliva specimens followed by pivotal CRISPR-Cas-assisted detection with lateral flow readout. The versatility of the developed platform can be implemented for CRISPR-Cas-based detections of other pathogens, showing great promise for POC diagnostics particularly in decentralized and resource-limited settings in low- and middle-income countries.

**Fig. 1.**
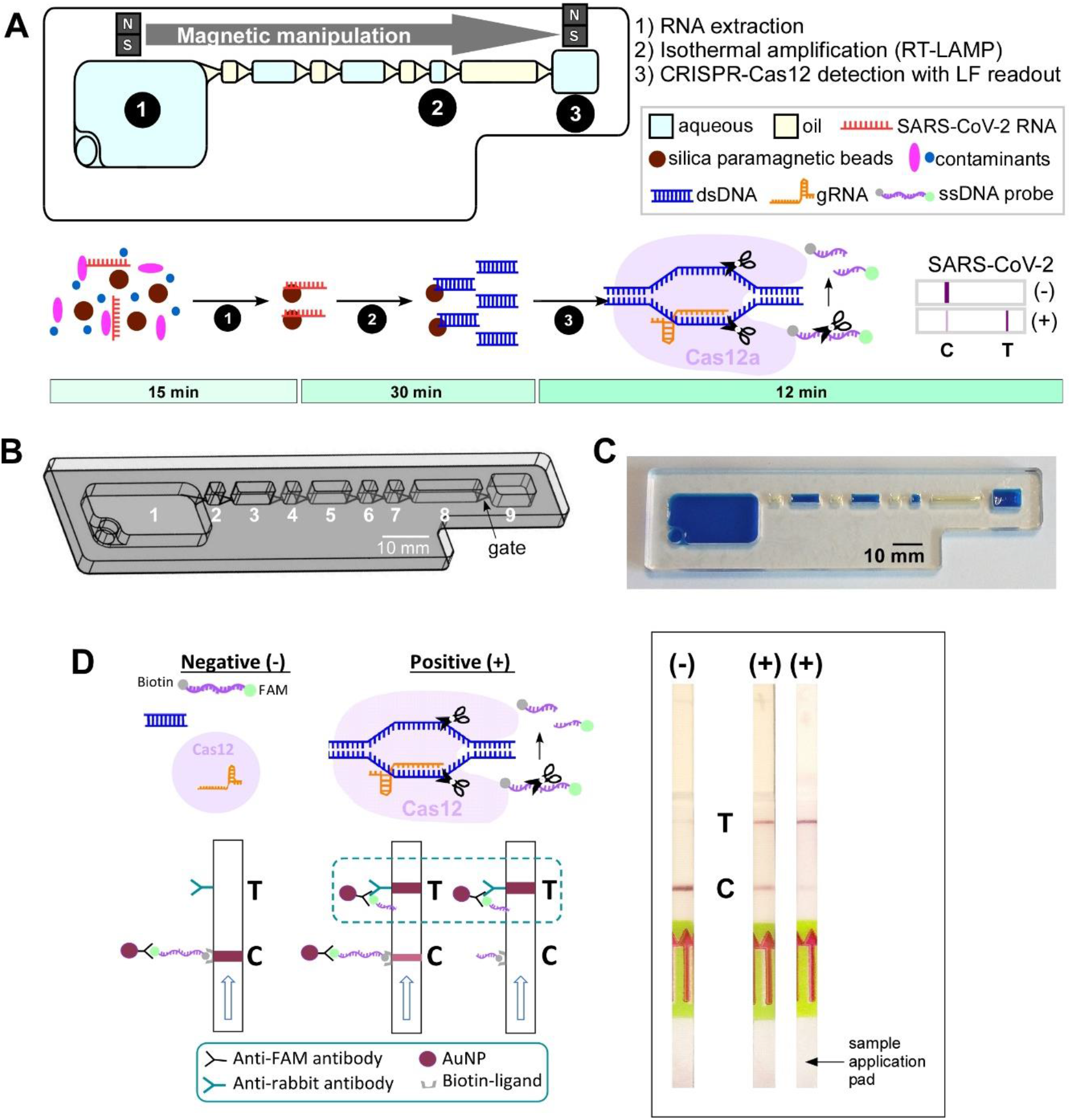
IFAST-CRISPR device for SARS-CoV-2 detection. (A) Design and (B) photograph of the IFAST-CRISPR device. Chamber 1 = sample + GuHCl + silica paramagnetic beads; chambers 2, 4, 6, 8 = mineral oil; chamber 7 = RT-LAMP reagent; chamber 9 = CRISPR-Cas12 reagent. (C) IFAST-CRISPR device detects SARS-CoV-2 viral RNA from unprocessed nasopharyngeal (NP) swab or saliva sample in a 1 h sample-to-answer workflow. Step 1: RNA is extracted from a sample via silica paramagnetic beads and 5 M GuHCl. Step 2: The MB-isolated RNA is *in vitro* transcribed and amplified into DNA amplicons via RT-LAMP. Step 3: The hybridization of targeted DNA sequence activates the gRNA-Cas12a complex to digest ssDNA probe, thereby producing a test line (**T**) on the lateral flow strip which can be visualized by the naked eye. (D) Principle of the lateral flow readouts for SARS-CoV-2 detection. Control line (**C**) appears from the intact FAM-biotinylated ssDNA reporter. Test line (**T**) is present from cleaved ssDNA reporter following target dsDNA-gRNA hybridization.

### Significance

Point-of-care COVID-19 diagnostics is critical to public health; however, current gold-standard RT-PCR methods are labour, equipment, and skill-intensive; resulting in far fewer tests being carried out in low- and middle-income countries such as Kenya. Affordable and highly reliable molecular testing using CRISPR-Cas-based assays with user-friendly sample collection and preparation would expand testing capacities and reduce community transmission in low-resource settings. Here, we describe an affordable sample-to-answer lab-on-a-chip platform which combines a simple sample preparation unit with highly-specific CRISPR-Cas assisted assays in one device, enabling 1 h detection of COVID-19 infections with a sensitivity comparable with gold standard laboratory-based methods.

## 2. Materials and Methods

### Ethical Approval

This study was approved by the Mount Kenya University Independent Ethical Review Committee (MKU/IERC/1811). Nasopharyngeal swab and saliva samples were collected from participants with their written informed consent after the nature and possible consequences of the study had been fully explained to them.

### Materials and reagents

Genomic SARS-CoV-2 RNA (2019-nCoV/USA-WA1/2020, ATCC VR-1986D), HCoV-OC43 (ATCC VR-1558D) and H1N1 (ATCC VR-1736D) RNAs were purchased from LGC standards, UK. SARS-CoV-2 verification panel (AccuPlex SARS-CoV-2, 0505-0168) was supplied by Technopath, UK. LAMP primers and lateral flow reporter were purchased from Integrated DNA Technologies (IDT). SYBR Safe, nuclease-free water, RNase decontamination solution and PCR adhesive film were supplied by Thermofisher Scientific, UK. Guanidine hydrochloride (GuHCl) and MagneSil paramagnetic beads were purchased from Promega. Lateral flow strips (Milenia HybriDetect 1) were purchased from TwistDx, UK.

### Device design, fabrication and preparation

The microfluidic device was made of polymethyl methacrylate and fabricated via CNC machine milling (Datron M7, Milton Keynes, UK). The device features a large sample chamber (1) (*w* = 15 mm, *l* = 26 mm); wash chambers (2), (4), (6) (*w* = 3 mm, *l* = 3 mm); wash chambers (3), (5) (*w* = 3 mm, *l* = 8.5 mm); RT-LAMP chamber (7) (*w* = 3 mm, *l* = 3 mm); chamber (8) (*w* = 3 mm, *l* = 15 mm); and a CRISPR-Cas detection chamber (9) (*w* = 6 mm, *l* = 9 mm). All chambers were 3.8 mm deep, and were interconnected via gates (*w* = 3 mm to 5 mm, *l* = 3 mm, *d* = 0.2 mm), Fig. 1B, C. The sample chamber contained a syringe port for a Luer fitting for sample introduction.

Devices were cleaned with RNase decontamination solution, followed by rinsing with nuclease-free water, and were left to dry at ambient temperature prior to use. The bottom of the device was sealed with PCR adhesive film.

### Tube-based RT-LAMP, followed by CRISPR-Cas12-assisted detection (DETECTR assays)

Tube-based DETECTR assays were performed using RT–LAMP for pre-amplification of genomic SARS-CoV-2 RNA using primers targeting N gene, and LbCas12a for the trans-cleavage assay following the protocol described by Broughton *et al*. (21) with some modifications. RT-LAMP was conducted at 64 °C for 30 min using 2 μL RNA templates serially diluted (10x) from 4,700 copies μL^-1^. RT-LAMP products were identified via agarose gel electrophoresis (1 %w/v agarose, 1xTAE buffer, 80 V, 45 min) and the results analyzed using a molecular imager (Chemidoc XRS+, BioRAD). LbCas12a-gRNA (RNP) complexes were generated prior to carrying out LbCas12 trans-cleavage assay, by pre-incubating LbCas12a (50 nM) with gRNA (62.5 nM) in 10xNEBuffer 2.1 at 37 °C for 30 min, followed by an addition of the lateral flow cleavage ssDNA reporter (/56-FAM/TTATTATT/3Bio/) to the reaction at a final concentration of 500 nM. CRISPR-Cas12-assisted detection was performed by mixing 2 μL of RT-LAMP amplicons with 22 μL of RNP complex containing lateral flow cleavage reporter and 76 μL of 1xNEBuffer 2.1, and incubating at 37 °C for 10 min. Subsequently, a lateral flow strip was added to the reaction tube and a result was visualized/photographed after ca. 2 min using a smartphone camera (SAMSUNG Galaxy A3). A negative sample was identified by the presence of a single band close to the sample pad (control line – C), whereas the presence of a single band close to the top of the test strip (test line – T), either with or without a control line, indicated a positive result (Fig. 1D). The gRNA specificity was evaluated against HCoV-OC43 and H1N1 RNAs using their respective primers (33).

### On-chip RNA extraction and RT-LAMP (Steps 1&2, Fig. 1A)

A sample with/without SARS-CoV-2 RNA was diluted in 5 M GuHCl containing 0.005% Tween 20 and added to the sample chamber (1) together with MagneSil paramagnetic beads (MBs, 1 μL). The mixture was gently agitated by hand or using a rotator set at 40 rpm (Stuart SB3, UK) for 5 min. Next, empty chambers were filled with mineral oil (chambers (2), (4), (6), (8)); RT-LAMP reagent overlaid with mineral oil (chamber (7)) and 0.005% Tween 20 (chambers (3), (5), (9)). The RNA-bound MBs were gathered, via the use of an external NdFeB magnet assembly (a 20 mm × 10 mm × 5 mm bar magnet and a 4 mm diameter × 5 mm height disc magnet; magnetic strength = 0.42 Tesla, Magnet Sales, UK) in the sample chamber and washed through wash liquids in chambers (2)-(6), and mixed with RT-LAMP reagent in chamber (7). The chip was subsequently placed in an incubator for RT-LAMP reaction at 64 °C for 30 min. The RT-LAMP amplicons (2 μL) were added into a tube containing CRISPR-Cas12 reagents (22 μL RNP complex containing lateral flow cleavage reporter and 76 μL 1xNEBuffer 2.1) and the reaction incubated at 37 °C for 10 min, prior to result visualization via lateral flow strip.

### On-chip CRISPR-Cas12 detection assays (Step 3, Fig. 1A)

The capability of the device for CRISPR-Cas12 detection assays was tested on tube-based RNA-extraction and RT-LAMP amplicons. MagneSil paramagnetic beads (1 μL) were added into a sample of genomic SARS-CoV-2 RNA diluted in 5 M GuHCl (940 copies mL^-1^) and mixed for 5 min by tube inversion. The RNA-extracted MB were washed with nuclease-free water (100 μL). The washed RNA-extracted MB were mixed with RT-LAMP reagent to undergo reaction at 64 °C for 30 min. The amplicons (2 μL) were transferred from the reaction tube into chamber (9) of the chip device which was prefilled with CRISPR-Cas12 assay mix and sealed with PCR adhesive film on the top to prevent contamination. Other chambers were also alternately filled with nuclease-free water (chambers (1), (3), (5), (7)), and mineral oil (chambers (2), (4), (6), (8)). On-chip CRISPR-Cas12 assays were performed at 37 °C for 10 min.

### Combined on-chip RNA extraction, RT-LAMP and CRISPR-Cas12-assisted detection (Steps 1-3, Fig. 1A)

On-chip RNA extraction and RT-LAMP were similarly conducted as described above. Extraction was performed for 5 min for samples containing free RNAs, and 15 min for SARS-CoV-2 verification panel samples. After performing RT-LAMP at 64 °C for 30 – 45 min, CRISPR-Cas12 reagent mix was added to chamber (9) and MBs from chamber (7) were moved into chamber (9) and the chamber was sealed with PCR adhesive film prior to reaction at 37 °C for 10 min and 2 min for lateral flow readout.

### Clinical sampling

Clinical samples from symptomatic COVID-19 infected patients (25-40 years old) confirmed by RT-PCR were retrieved from a national testing centre in Kenya Medical Research Institute. The sampling was authorized by the Ethical Review Committee of Mount Kenya University (MKU/IERC/1811) and carried out in accordance with the approved guidelines.

### Cigarette filter as saliva collection material

A Swan brand cigarette filter was placed inside a 1 mL disposable syringe (BD Plastipak) prior to drawing a sample containing 4,700 copies mL^-1^ genomic SARS-CoV-2 until the sample (ca. 0.2 mL) filled the filter (Fig. S2, Supporting Information). The end of the syringe was subsequently placed into a container of 5 M GuHCl solution where drawing of GuHCl through the cigarette filter was conducted until the liquid level reached the 1 mL mark. Next, the syringe was fitted into the syringe holder in the sample chamber of the IFAST-CRISPR device and the liquid inside was expelled into the sample chamber using the syringe plunger. Further on-chip steps for RNA extraction, amplification and CRISPR-Cas12 detection were carried out following the protocols described above.

The Swan brand cigarette filters were additionally tested with dilutions of a saliva samples spiked with SARS-CoV-2 positive nasopharyngeal swab samples. Several 10-fold serial dilutions of the clinical RT-PCR-confirmed positive nasopharyngeal swab samples were prepared using saliva donated by a COVID-19 negative donor. The diluted samples (600 μL) were aliquoted into Eppendorf tubes, into which the filters were immersed. The content was transferred into a new Eppendorf tube by squeezing the filters using tweezers and the volume transferred adjusted to 150 μL for all the dilutions. As a control, 150 μL of the diluted samples were taken for each dilution and detection of COVID-19 was performed in pairs. RNA extraction was carried out using Ribo-Virus RNA extraction kit (Sacace Biotechnologies) and RT-PCR conducted using a TBG Q6000 real time PCR system (TBG Biotechnology).

## 3. Results and Discussion

### 3.1 Development of IFAST-CRISPR microfluidic device for SARS-CoV-2 detection in resource-limited settings

The IFAST-CRISPR device (Fig. 1B, C) has been designed to combine streamlined IFAST sample preparation for rapid and effective RNA extraction from viral particles, with sensitive and highly specific CRISPR-Cas-associated detection. The design was adapted from our previously reported IFAST RT-LAMP device (2); however, in this instance, the chambers were horizontally aligned for easier magnetic manipulation, multiplexing and automation (Fig. 1C); as opposed to the meandering chamber layout in the previous design which was chosen in order to minimise carry-over and device footprint. The gates and interconnecting chambers were of similar dimensions to those of the previous design (Fig. 1B); permitting up to 60 min heating at 65 °C with intact immiscible interfaces (2). The operation steps for SARS-CoV-2 detection utilizing the IFAST-CRISPR device are (i) loading the sample chamber with viral sample diluted in 5 M GuHCl and incubating the sample mix with silica paramagnetic beads either by manual agitation or using a shaking/rotating platform, (ii) adding oil, washing liquid and RT-LAMP reagent, (iii) isolating the magnetic beads from the sample and washing through immiscible liquids using magnetic manipulation across the bottom of the device, to allow washed magnetic beads to mix with RT-LAMP reagents, (iv) performing RT-LAMP by placing the device in an incubator or over a hot plate, (v) adding CRISPR-Cas12a reagent in the last chamber and move the magnetic beads from RT-LAMP chamber into CRISPR-Cas12 detection chamber, (vi) carrying out CRISPR-Cas12a detection in an incubator/on a hot plate, (vii) adding a lateral flow test strip into the CRISPR-Cas12a chamber for result visualization.

The platform simplified the cumbersome RNA extraction process normally required prior to performing CRISPR-Cas-based assays for SARS-CoV-2 detection (4). This was achieved by combining the lysis and magnetic bead-binding steps, and eliminating the ethanol wash and elution steps typically conducted in multi-step solid phase RNA extraction (3). The duration of the sample extraction time was reduced to just 15 min with minimal hands-on time. The relatively large sample chamber accommodating up to 1.5 mL sample, coupled with the concentration of analyte by magnetic beads, will benefit the detection of low viral loads. MagneSil paramagnetic beads were chosen over oligo (dT)-functionalized magnetic beads employed in the previous IFAST RT-LAMP device (2), in order to reduce the cost per assay (Table S1, Supporting Information). Additionally, MagneSil beads can be stored at room temperature (15 – 30 °C), making them more suitable for limited-resource settings than oligo (dT)-functionalized beads which require a cold chain storage (2 – 8°C). The chaotropic salt GuHCl was employed in the lysis and binding step as well as acting as RNase inhibitor to help maintain RNA integrity in clinical samples. Inexpensive GuHCl salt can be pre-stored inside the device in a solid form, or directly used as GuHCl (aq) for sample collection instead of universal/viral transport media (UTM/VTM), avoiding supply shortage of UTM/VTM during periods of high demand.

By passing RNA-bound MBs through two sets of immiscible wash liquids, rapid purification of the magnetic bead-isolated RNA was achieved within a few seconds, thereby protecting the RNA integrity for the subsequent amplification and detection steps.

The following amplification and detection steps were adapted from the reported SARS-CoV-2 DETECTR protocol from Broughton *et al*. (21) where simultaneous reverse transcription and isothermal amplification were achieved by RT-LAMP, followed by CRISPR-Cas12 detection of target DNA amplicons. As well as being affordable and widely accessible, the reported LAMP N gene primer set was also granted Emergency Use Authorization (EUA) by the U.S. Food and Drug Administration for Mammoth Biosciences (40). Despite being active at 16 – 48 °C, LbCas12a(Cpf1) is commercially available and more accessible than the thermostable AapCas12b exploited in the STOPCovid system (28). Tube-based CRISPR-Cas assays require the transfer of minute quantities of DNA target from the amplification tube into the CRISPR-Cas detection tube (5, 6). By including an additional oil chamber after the RT-LAMP chamber on the IFAST-CRISPR device, magnetic manipulation of amplicons-bound on MBs into the CRISPR-Cas detection chamber can easily be achieved, preventing potential contamination introduced by amplicon transfer by pipetting.

A lateral flow test strip was used for result interpretation instead of fluorescence detection to avoid utilizing a complex equipment set up. The test strips embedded biotin-ligand and anti-FITC/FAM antibodies in the control and test lines, respectively (Fig. 1D). The ssDNA lateral flow reporter was labelled on the 5’-end with FAM and on the 3’-end with biotin. If no target DNA is present (SARS-CoV-2 negative sample), ssDNA reporters remain intact, resulting in a strong control line close to the sample application pad. While on the contrary, positive samples containing target DNA which when hybridized with the gRNA, will trigger collateral cleavage of the ssDNA reporters in the proximity, activating a test line closer to the end of the test strip. This unambiguous detection step takes 1-2 min, and results can be analyzed by the naked eye, facilitating device deployment in resource-poor settings where readily and accurately interpreted results are necessary.

### 3.2 CRISPR-Cas-assisted detection of SARS-CoV-2

Validation of the CRISPR-Cas12a-based assays for SARS-CoV-2 detection was first carried out in tubes on a series of ten-fold dilutions of genomic RNA with initial concentration of 4700 copies μL^-1^. Utilising primers targeting N gene, positive amplifications were observed from 4.7 copies of genomic RNA after 30 min (Fig. 2a, gel electrophoresis); a much higher sensitivity compared with the pH-dependent colorimetric RT-LAMP assay reported in our previous study where ≥ 470 genome copies were amplified at the same reaction time (33). Visual signals observed from 10 min CRISPR-Cas12 detection in the following step confirmed the sensitivity of the assay to detect down to 4.7 genome copies (Fig. 2A, lateral flow strips). The specificity of the gRNA in the CRISPR-Cas12 assay only towards SARS-CoV-2 with no cross-reactivity with the related HCoV-OC43 and influenza A H1N1 is demonstrated in Fig. 2B. Although positive amplifications were detected with correct pairing between RNAs and their respective primers (Fig. 2B, gel electrophoresis), a test line only appeared on the strip from the SARS-CoV-2 sample as a result of Cas12 trans-cleavage activity following hybridization of the gRNA-targeted DNA sequence.

**Fig. 2.**
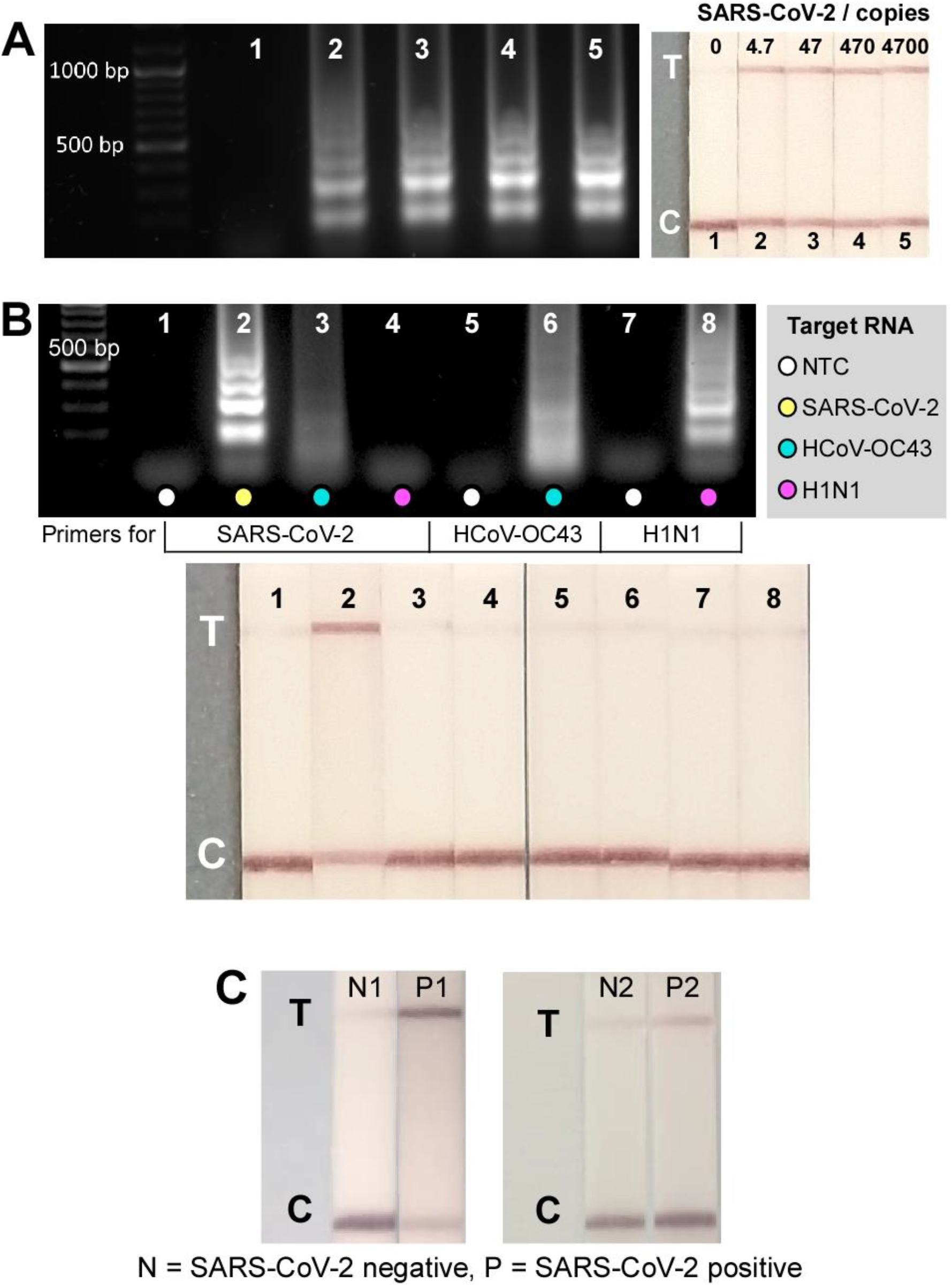
Tube-based DETECTR assays. (A) Sensitivity test of tube-based DETECTR assays on genomic SARS-CoV-2 RNA. Left: Gel electrophoresis results from RT-LAMP products after 30 min reaction at 64 °C. Right: Lateral flow strips from CRISPR-Cas12a detection of RT-LAMP products from left, demonstrating a detection of 4.7 genome copies in 50 min (30 min RT-LAMP + 20 min CRISPR-Cas12a detection; n=2). (B) Specificity test of CRISPR-Cas12a assay – test line only appeared on SARS-CoV-2 strip suggesting gRNA specificity only to SARS-CoV-2 with no cross-reactivity towards HCoV-OC43 and H1N1, although positive amplifications were observed in such RNAs with their respective primers after RT-LAMP (gel). Reactions performed using 5 pg HCoV-OC43, 28.9 pg H1N1, and 95 pg SARS-CoV-2 (n=1). (C) Lateral flow test strips from tube-based DETECTR assays from negative samples (N1&N2) and SARS-CoV-2 RNA extracted from nasopharyngeal samples (P1&P2).

The CRISPR-Cas-assisted assay was further tested with RNA extracted from nasopharyngeal swab samples (Fig. 2C); demonstrating successful detection of SARS-CoV-2 RNA in RT-qPCR confirmed positive samples (strips 2 and 4 with Ct values of 21 and 18, respectively). Notably, the positive readout signal from tube-based DETECTR assay that was performed on sample 4 agreed with the RT-qPCR result and confirmed its superior sensitivity over the antigen lateral flow assay where negative result was observed.

### 3.3 Evaluation of the IFAST-CRISPR device for SARS-CoV-2 detection

In order to apply the IFAST-CRISPR device for SARS-CoV-2 detection, individual steps on-chip were systematically evaluated. Utilizing silica paramagnetic beads combined with guanidinium salt, genomic SARS-CoV-2 was extracted and purified through the immiscible liquids, and finally underwent RT-LAMP on chip. The amplification products were subsequently detected via tube-based CRISPR-Cas12 assay. Verification of target DNA amplicons successfully generated from magnetically-isolated RNA from initial samples containing ≥ 470 genome copies mL^-1^ is demonstrated in Fig. 3A.

**Fig. 3.**
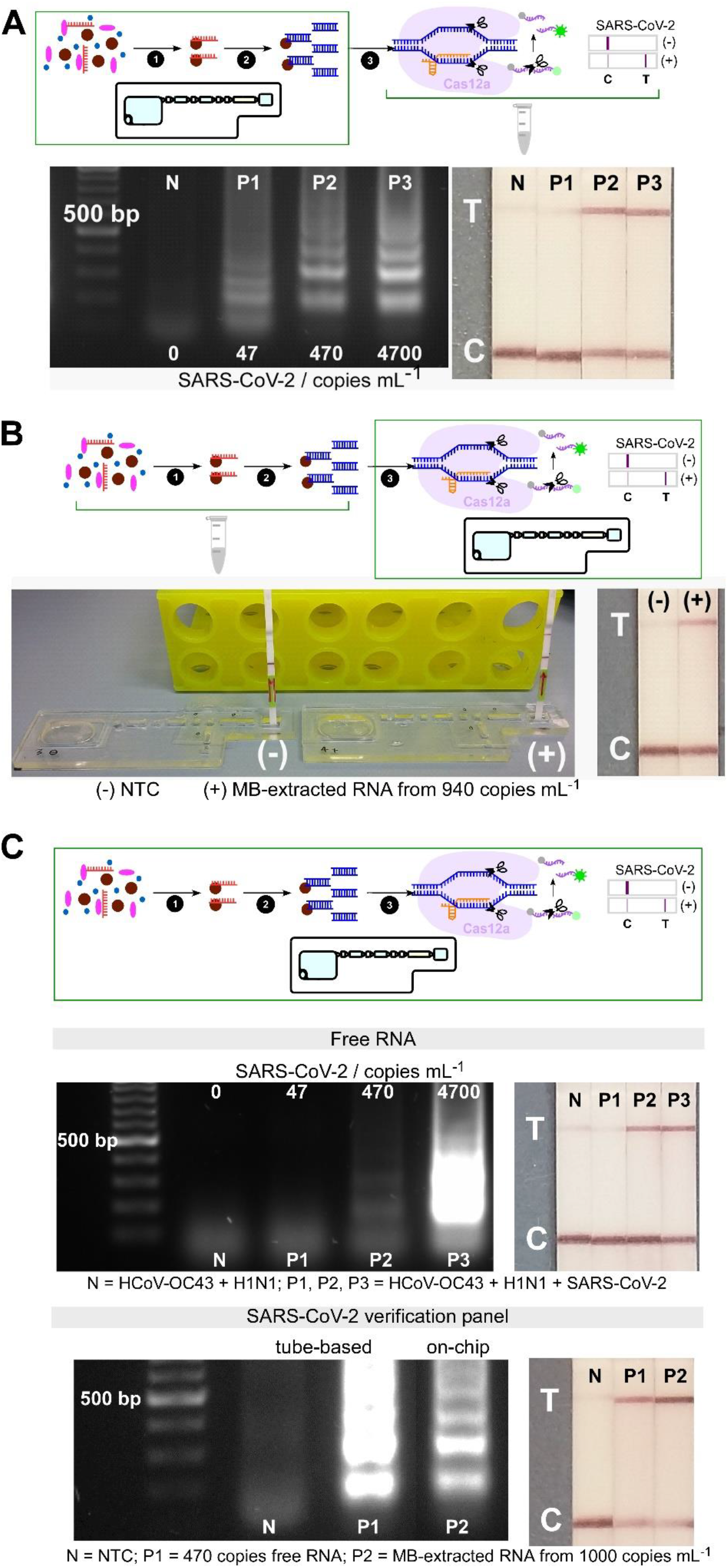
Analytical validation of individual and combined on-chip assays. (A) On-chip RNA extraction, followed by RT-LAMP assays; gel electrophoresis results showing target dsDNA being amplified from MB-extracted RNA from ≥ 470 copies mL^-1^ initial concentrations, confirmed by test lines on lateral flow test strips (n=2). (B) On-chip CRISPR-Cas12 assays of amplicons from tube-based RNA extraction and RT-LAMP - collateral cleavage of lateral flow ssDNA reporters following the hybridization between the gRNA and dsDNA target showing a test line in positive sample from MB-extracted RNA (n=1). (C) On-chip integrated steps of RNA extraction, RT-LAMP and CRISPR-Cas-assisted detection from samples containing free genomic SARS-CoV-2 RNA (in a mixture containing HCoV-OC43 and H1N1 RNAs, n=2), and from viral particles containing SARS-CoV-2 genome (SARS-CoV-2 verification panel, n=1).

The feasibility of performing a CRISPR-Cas12a assay for the specific detection of target DNA on-chip was next investigated. The corresponding readout signals from negative and positive samples suggested the viability of the IFAST-CRISPR device as a reactor for the trans-cleavage activity of Cas12a to detect target amplicons (Fig. 3B).

The performance of the IFAST-CRISPR device was next examined on the combined workflow for extraction, RT-LAMP and CRISPR-Cas12a detection of samples containing genomic RNA (Fig. 3c – top). Negative sample containing HCoV-OC43 and H1N1 RNAs showed only a control line on the lateral flow test strip, whilst test lines were present from positive samples containing HCoV-OC43, H1N1 and ≥470 copies mL^-1^ SARS-CoV-2 RNAs. These results not only validated the successful integration of consecutive steps on-chip, but they also confirmed the assay specificity as previously described in Fig. 2b. The assay performance of the IFAST-CRISPR device was further evaluated using replication-deficient SARS-CoV-2 viral particle samples (SeraCare AccuPlex SARS-CoV-2 Verification Panel). By mixing the viral sample with silica paramagnetic beads and 5 M GuHCl, lysis of viral particles and binding of SARS-CoV-2 RNA to magnetic beads were accomplished simultaneously (Fig. S2). Translating the same process onto the IFAST-CRISPR device, coupled with rapid washing through minute quantities of immiscible liquids, demonstrated that RNA can be magnetically isolated and detected from contrived viral samples containing 1,000 genome copies mL^-1^ in 15 min (Fig. 3C – bottom). RT-LAMP DNA amplicons generated from the magnetically-isolated RNA, verified by gel electrophoresis, confirmed the efficient on-chip purification process in eliminating possible RT-LAMP assay inhibitor crossing over from the sample chamber. Finally, recognition of target DNA sequences that had been magnetically transported to the CRISPR-Cas12a detection chamber was performed. This was achieved by using the Cas12a gRNA initiated collateral cleavage of ssDNA reporter, leading to a test line on the lateral flow strip, similar to the tube-based assay for positive control of free SARS-CoV-2 RNA. This simple, and yet complete on-chip workflow has already shown a similar level of detection as the gold standard RT-qPCR (1,000 copies mL^-1^) and has the potential to be further optimized.

### 3.4 Easy access and cost-effective cigarette filters for saliva collection

Another aim of the present study was to replace the uncomfortable nasopharyngeal (NP) or oropharyngeal (OP) swabbing with saliva sampling using a cigarette-type filter. Although NP and OP swab specimens have been considered reliable and are most often used for COVID-19 diagnostics (7), (6), acquiring an NP/OP swab can be unpleasant, and can initiate coughing leading to increasing risk of nosocomial spread of respiratory viruses (41), as well as in suboptimal specimens being collected dependent upon the experience of the collector. Being inexpensive and easy to access, Swan brand cigarette filters were investigated as potential substrate for saliva collection for COVID-19 screening in low-resource settings. The feasibility of introducing a viral sample from a Swan cigarette filter for the IFAST-CRISPR device was examined using genomic SARS-CoV-2 RNA (Fig. S2 and Fig. 4A). No adverse effect from the filters was observed either on RT-LAMP or CRISPR-Cas12a assays performed on-chip. Additionally, Swan brand filters were tested with dilutions of a SARS-CoV-2 positive nasopharyngeal swab specimens. After immersing a filter in 1 mL of each dilution, the liquid retained within the filters was extracted and subjected to RT-qPCR. Ct values obtained from the samples retained within the filters compared favourably to those of the initial samples without filters, indicating neither RT-qPCR assay inhibition nor compromised RNA integrity by the use of Swan filters (Fig. 4b). These proof-of-concept methodological approaches verify the potential use of Swan filters for saliva collection, which can be directly interfaced with the IFAST-CRISPR device.

**Fig. 4.**
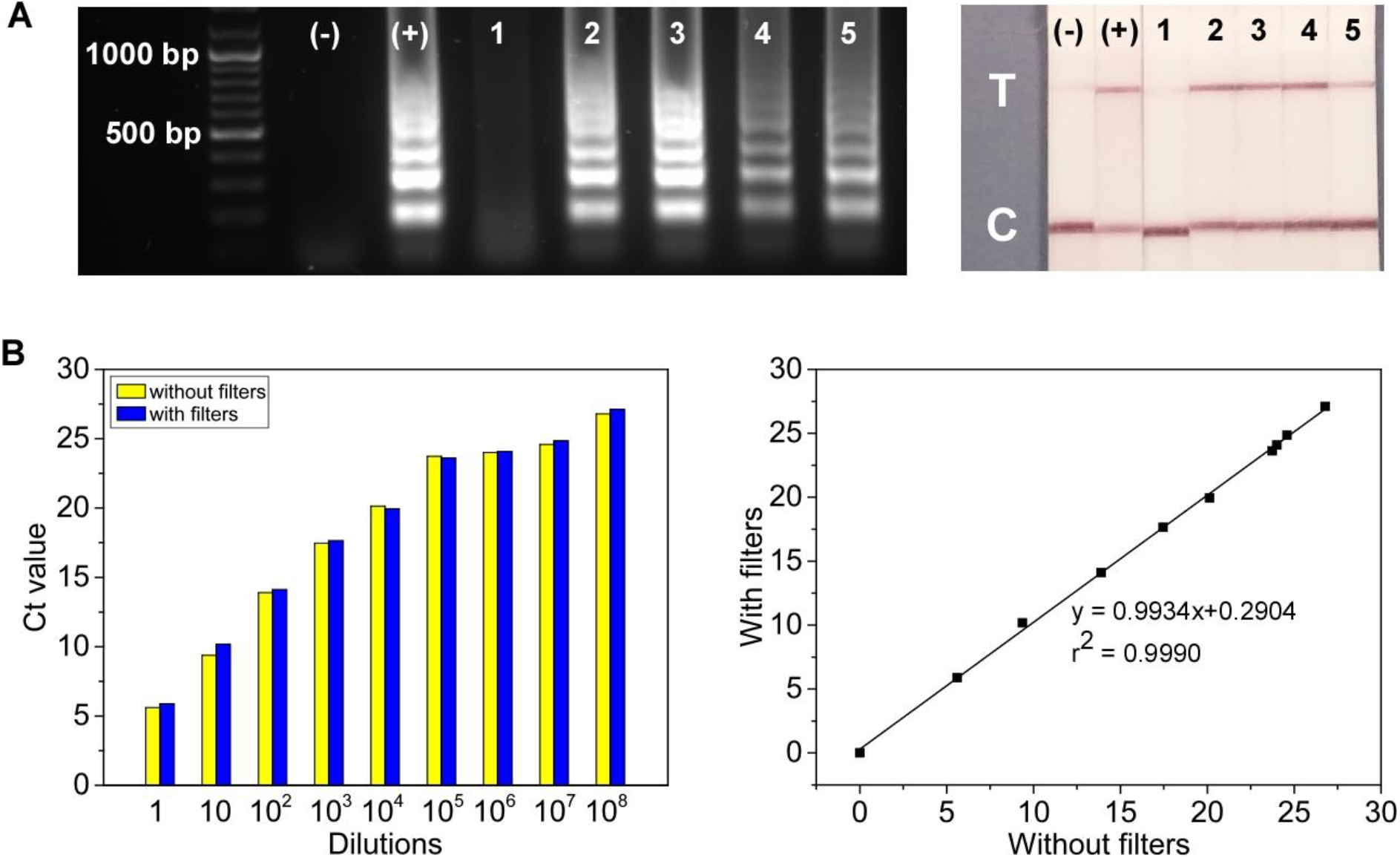
Swan brand filters tested with genomic SARS-CoV-2 RNA and clinical samples. (A) Gel electrophoresis and lateral flow strips from RT-LAMP assays and CRISPR-Cas12a detection, respectively. Control experiments without cigarette filters: (-) = No template control; (+) = 470 copies of free SARS-CoV-2 RNA. Samples collected using cigarette filters: tube-based assays; 1 = nuclease-free water; 2 = initial sample containing 94 genome copies; 3 = sample extracted from cigarette filter immersed in 47 copies μL^-1^ of RNA sample. On-chip assays; 4, 5 = magnetic bead-extracted RNA extracted from cigarette filters with initial concentration of 4700 copies mL^-1^. (B) Ct values of samples before and after being subjected to Swan filters from various dilutions of a SARS-CoV-2 positive saliva samples (n = 1).

The present manuscript demonstrated the manual operation of an IFAST-CRISPR device, for use as a molecular COVID-19 diagnostic tool, by a semi-trained operator in resource-limited laboratories in sub-Saharan Africa, *e*.*g*., Kenya. The device has been designed to operate with already available laboratory facilities such as incubators for heating and shakers for RNA extraction. Although presently being fabricated by CNC milling, the thermoplastic polymethyl methacrylate devices can be mass produced by injection moulding. The total cost of a device including materials and reagents is currently around 8 USD (Table S1, Supporting Information).

## Conclusions

We have demonstrated a microfluidic platform which integrated all essential steps necessary for SARS-CoV-2 detection: as an affordable COVID-19 molecular diagnostic device with similar sensitivity and specificity to the gold standard RT-qPCR tests. The platform required only a basic heating source such as simple incubators or hot plates which are usually available in most laboratories in low-resource settings, without the need for costly or specialized instruments. This simple, and yet highly versatile platform can potentially be applied not only for COVID-19 screening, but also for other pathogenic infections. This allows affordable, sensitive, target-specific, quick turnaround nucleic amplification tests (NAATs) to be achieved where rapid and precise diagnostic tools are needed for infection containment as well as for timely treatment.

## Supporting information

supporting information to be used for preprint BIORXIV-2021-474287 - Version 1

## Data Availability

All data produced in the present work are contained in the manuscript.

## Acknowledgments

The study was financially supported by the Newton-Utafiti Fund Kenya Country Prize 2020.

