## supporting information to be used for preprint BIORXIV-2021-474287 - Version 1 for "An integrated lab-on-a-chip device for RNA extraction, amplification and CRISPR-Cas12a-assisted detection for COVID-19 screening in resource-limited settings"

**RNA extraction from viral particles using silica paramagnetic beads and GuHCl solution**

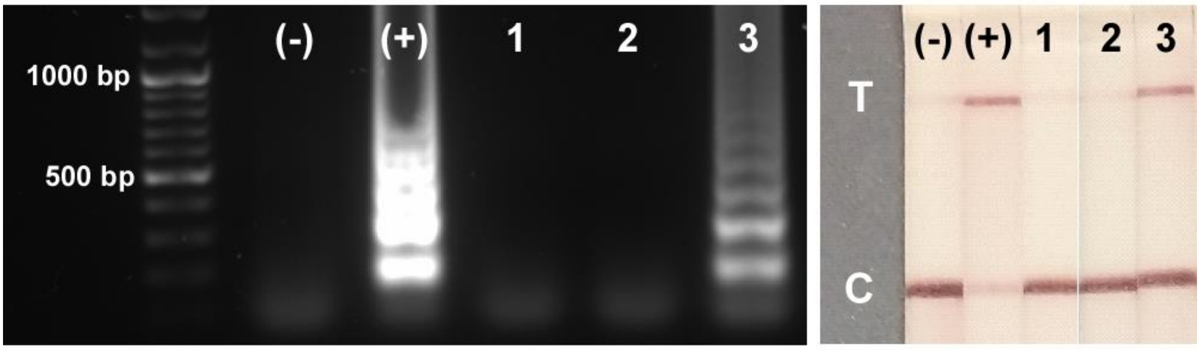


**Fig. S1** Gel electrophoresis of tube-based RT-LAMP, and lateral flow test strips from CRISPR-Cas12a detection of RT-LAMP products. Samples: (-) = NTC; (+) = 470 copies of genomic SARS-CoV-2 (free RNA); 1 = 100 copies $mL^{-1}$ of SARS-CoV-2 verification panel; 2 = MB + SARS-CoV-2 verification panel in nuclease-free water (1000 copies $mL^{-1}$); 3 = MB + SARS-CoV-2 verification panel in 5 M GuHCl (1,000 copies $mL^{-1}$). Incubation time for MB and SARS-CoV-2 verification panel = 10 min. RT-LAMP was performed at 64 °C for 35 min using primers targeting N gene. CRISPR-Cas12a assays were carried out at 37 °C for 10 min.

**Cigarette filter coupled with IFAST-CRISPR/Cas device for saliva collection and SARS-CoV-2 detection**

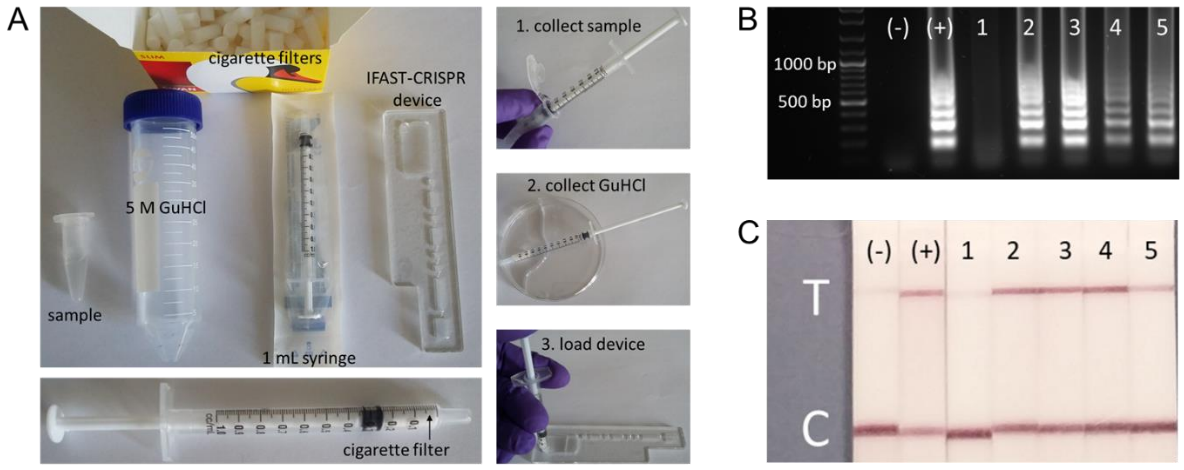


**Fig. S2** A) Interfacing of Swan brand cigarette filter for sample collection and IFAST-CRISPR on-chip assays. B) Gel electrophoresis of tube-based RT-LAMP, and C) lateral flow test strips from CRISPR-Cas12a detection of RT-LAMP products. Samples: (-) = NTC; (+) = 470 copies of genomic SARS-CoV-2 (free RNA); 1 = 100 copies $mL^{-1}$ of SARS-CoV-2 verification panel; 2 = MB + SARS-CoV-2 verification panel in nuclease-free water (1,000 copies $mL^{-1}$); 3 = MB + SARS-CoV-2 verification panel in 5 M GuHCl (1,000 copies $mL^{-1}$). Incubation time for MB and SARS-CoV-2 verification panel = 10 min. RT-LAMP was performed at 64 °C for 35 min using primers targeting N gene. CRISPR-Cas12a assays were carried out at 37 °C for 10 min.

**Table S1.** Cost breakdown for device and reagents.

| | Component | Supplier | Amount per purchase unit | | Required amount per device/assay | | Number of device/assay per unit | Unit price (USD) | Price per device/assay (USD) |
|---|---|---|---|---|---|---|---|---|---|
| | | | value | unit | value | unit | | | |
| Device materials | PMMA device | Kingston Plastics | 1 | sheet | 0.05 | sheet | 20 | 4.70 | 0.24 |
| | Adhesive PCR plate seals | Thermofisher Scientific | 100 | sheet | 0.5 | sheet | 200 | 115.73 | 0.58 |
| Reagents | Magnesil Beads | Promega | 25 | mL | 0.001 | mL | 25,000 | 223.49 | 0.01 |
| | GuHCl | Promega | 100 | g | 0.478 | g | 209.36 | 61.91 | 0.30 |
| | Milenia HybriDetect 1 Lateral flow strips | TwistDx | 100 | strip | 1 | strip | 100 | 296.26 | 2.96 |
| | LbCas12a | New England Biolabs | 70 | µL | 1 | µL | 70 | 87.46 | 1.25 |
| | gRNA – N gene | Eurofins | 122,048 | µL | 1.25 | µL | 152,560 | 603.01 | 0.004 |
| | gRNA – E gene | Eurofins | 122,048 | µL | 1.25 | µL | 152,560 | 262.28 | 0.0017 |
| | gRNA – RNase P | Eurofins | 122,048 | µL | 1.25 | µL | 152,560 | 262.28 | 0.0017 |
| | LF reporter | Integrated DNA Technologies | 2107 | µL | 0.5 | µL | 4,214 | 79.84 | 0.02 |
| | LAMP primers – N gene FIP | Integrated DNA Technologies | 931 | µL | 0.4 | µL | 2,328 | 4.96 | 0.002 |
| | LAMP primers – N gene BIP | Integrated DNA Technologies | 956 | µL | 0.4 | µL | 2,390 | 4.72 | 0.002 |
| | LAMP primers – N gene F3 | Integrated DNA Technologies | 10,250 | µL | 0.05 | µL | 205,000 | 2.18 | 1.06E-05 |
| | LAMP primers – N gene B3 | Integrated DNA Technologies | 11,000 | µL | 0.05 | µL | 220,000 | 2.66 | 1.21E-05 |
| | LAMP primers – N gene LF | Integrated DNA Technologies | 3,675 | µL | 0.1 | µL | 36,750 | 2.3 | 6.26E-05 |
| | LAMP primers – N gene LB | Integrated DNA Technologies | 4,525 | µL | 0.1 | µL | 45,250 | 2.18 | 4.82E-05 |
| | LAMP primers – E gene FIP | Integrated DNA Technologies | 931 | µL | 0.4 | µL | 2,328 | 4.96 | 0.002 |
| | LAMP primers – E gene BIP | Integrated DNA Technologies | 956 | µL | 0.4 | µL | 2,390 | 4.72 | 0.002 |
| | LAMP primers – E gene F3 | Integrated DNA Technologies | 10,250 | µL | 0.05 | µL | 205,000 | 2.18 | 1.06E-05 |
| | LAMP primers – E gene B3 | Integrated DNA Technologies | 11,000 | µL | 0.05 | µL | 220,000 | 2.66 | 1.21E-05 |

| | | | | | | | | | |
|---|---|---|---|---|---|---|---|---|---|
| | LAMP primers – E gene LF | Integrated DNA Technologies | 3,675 | µL | 0.1 | µL | 36,750 | 2.3 | 6.26E-05 |
| | LAMP primers – E gene LB | Integrated DNA Technologies | 4,525 | µL | 0.1 | µL | 45,250 | 2.18 | 4.82E-05 |
| | LAMP primers – RNase P - FIP | Integrated DNA Technologies | 931 | µL | 0.4 | µL | 2,328 | 4.96 | 0.002 |
| | LAMP primers – RNase P - BIP | Integrated DNA Technologies | 956 | µL | 0.4 | µL | 2,390 | 4.72 | 0.002 |
| | LAMP primers – RNase P – F3 | Integrated DNA Technologies | 10,250 | µL | 0.05 | µL | 205,000 | 2.18 | 1.06E-05 |
| | LAMP primers – RNase P – B3 | Integrated DNA Technologies | 11,000 | µL | 0.05 | µL | 220,000 | 2.66 | 1.21E-05 |
| | LAMP primers – RNase P - LF | Integrated DNA Technologies | 3,675 | µL | 0.1 | µL | 36,750 | 2.3 | 6.26E-05 |
| | LAMP primers – RNase P - LB | Integrated DNA Technologies | 4,525 | µL | 0.1 | µL | 45,250 | 2.18 | 4.82E-05 |
| | Bst 2.0 DNA polymerase | New England Biolabs | 200 | µL | 1 | µL | 200 | 96.85 | 0.48 |
| | Warmstart RTx Reverse Transcriptase | New England Biolabs | 25 | µL | 0.5 | µL | 50 | 87.46 | 1.75 |
| | dNTP mix | New England Biolabs | 800 | µL | 3.5 | µL | 228.57 | 87.46 | 0.38 |
| | | | | | | | | **Total** | **7.99** |
